# PANGENOMES AID ACCURATE DETECTION OF LARGE INSERTION AND DELETIONS FROM GENE PANEL DATA: THE CASE OF CARDIOMYOPATHIES

**DOI:** 10.1101/2024.11.27.24318059

**Authors:** Francesco Mazzarotto, Özem Kalay, Elif Arslan, Valeria Cinquina, Deniz Turgut, Rachel J Buchan, Mona Allouba, Valeria Bertini, Sarah Halawa, Pantazis Theotokis, Gungor Budak, Francesca Girolami, Petra Peldova, Jiri Bonaventura, Yasmine Aguib, Marina Colombi, Iacopo Olivotto, Massimo Gennarelli, Milan Macek, Elisabetta Pelo, Marco Ritelli, Magdi Yacoub, Paul JR Barton, H Serhat Tetikol, Roddy Walsh, James S Ware, Amit Jain

## Abstract

Gene panels represent a widely used strategy for genetic testing in a vast range of Mendelian disorders. While this approach aids reliable bioinformatic detection of short coding variants, it fails to detect most larger variants. Recent studies have recommended the adoption of pangenomes to augment detection of large variants from targeted sequencing, potentially providing diagnostic laboratories with the possibility to streamline diagnostic work-ups and reduce costs.

Here, we analyze a large-scale cohort comprising 1,952 cardiomyopathy cases and 1,805 technically matched controls and show that a pangenome-based workflow, GRAF, conjugates higher precision and recall (F1 score 0.86) compared with conventional orthogonal methods (F1 0-0.57) in detecting potentially pathogenic ≥20bp variants from short-read panel data.

Our results indicate that pangenome-based workflows aid precise and cost-effective detection of large variants from targeted sequencing data in the clinical context. This will be particularly relevant for conditions in which these variants explain a high proportion of the disease burden.

## 1. INTRODUCTION

Large sequence changes, such as copy number variants, have historically been difficult to detect from short-read targeted sequencing assays like whole-exome sequencing (WES) or gene panels, due to limitations intrinsic to these technologies^1,2^. This is a relevant drawback in the clinical context, as – in spite of decreasing costs of short- and long-read whole-genome sequencing (WGS) – short-read (usually 150bp) WES and targeted gene panels still represent by far the most widely used approaches for the genetic diagnosis of inherited conditions.

Routine adoption of WGS, especially if based on long-read sequencing, would streamline detection of large variants and would undoubtedly carry several other advantages (such as the possibility to detect pathogenic variants in the non-coding regions of the genome), but targeted sequencing approaches, such as gene panels, are still characterized by markedly lower costs accompanied by diagnostic yields often not significantly different from those characterizing WGS^3–6^.

Other potentially cost-effective and high-throughput approaches such as optical genome mapping are still not widespread or approved for diagnostics, and clinical laboratories still rely on wet lab technologies such as MLPA and array CGH to detect large sequence variants. While these approaches enable reliable detection of large duplications and deletions, they are characterized by low throughput and high costs, besides being laboratory intensive. Furthermore, these strategies don’t generally enable breakpoint resolution, not being informative on the exact locations where variants occur.

For these reasons, the availability of computational approaches able to detect large variants in a reliable and efficient manner from short-read targeted sequencing assays would be extremely advantageous for diagnostic laboratories, providing the possibility to reduce costs while also streamlining large variant identification. Many tools, achieving a wide range of performances in terms of precision and recall, have been developed for this purpose^7,8^. The underlying computational approaches used for variant detection are generally divided into 4 categories, depending on whether the algorithm relies on changes in read depth, on the presence of split reads, on the identification of abnormal mapping distances between paired reads, or on de novo assembly. Each of these approaches is characterized by advantages and drawbacks, with several tools exploiting multiple types of signals^9^. Read depth-based approaches, for example, are adopted by most tools and are particularly suitable to estimate the precise copy number when detecting duplications and deletions. However, these are blind towards other classes of variants such as inversions, and often require at least one control sample to use as reference. De novo assembly-based methods, on the contrary, are robust toward inaccurate read mapping and enable base-pair breakpoint resolution but are adopted by a small proportion of tools as they are computationally overwhelming and slow.

The differential advantages and downsides characterizing virtually every specific tool served as foundation for the conceptualization of pipelines combining results from multiple large variant callers to increase accuracy^10–12^. However, the routine application of bioinformatic pipelines entailing the usage of multiple tools can represent a hurdle in diagnostic laboratories without a dedicated bioinformatics team.

In this context, the advent of pangenome references (originally called graph reference genomes), as opposed to the single haplotype reference assemblies such as GRCh38, represented the beginning of a revolution as far as large variant detection from short-read targeted sequencing is concerned.

Pangenome references include the conventional reference genome as well as known genetic variation and alternative haplotypes, obtained from databases and cohorts^13^. This not only makes them variant-aware, with a consequent significant improvement of read mapping accuracy, but also renders them tailorable, for example to a given ancestry^14^. The augmented accuracy in read alignment translates into substantial improvements in variant identification, with this being especially pronounced for large variants^15^. As a result, the adoption of a pangenome reference has been shown to augment the precise identification of large genetic variants in plants as well as in human Mendelian disease, across both short- and long-read sequencing^16,17^.

However, the impact of the routine adoption of a pangenome-based approach in the clinical context has not been assessed to date. Here, we analyzed a multi-centric cohort of 1969 cardiomyopathy cases and 1805 controls sequenced with the Illumina Trusight Cardio panel^18^ – widely used in the diagnostic context – with the Velsera pangenome-based GRAF pipeline^19^ as well as three other orthogonal approaches to detect variants larger than 20bp, selected for their specific strengths for analyzing this type of data.

While large variants are known to be minor contributors to the cardiomyopathy disease burden^20–22^, our performance comparison between GRAF and multiple other tools in detecting large sequence variants from short-read targeted sequencing data provides the scientific community with findings applicable to any condition for which targeted sequencing panels are a widely adopted genetic testing strategy. Results show GRAF to conjugate the highest recall with the absence of false positive calls and suggest how – with further refinements – pangenome-based approaches may soon yield comparable performances to those characterizing wet lab-based techniques in detecting large variants. This would contribute to a striking cost reduction for diagnostic laboratories, especially concerning conditions where large variants explain large proportions of the disease burden.

## 2. MATERIALS AND METHODS

### 2.1 Study cohorts

A total of 3774 samples sequenced on the Illumina Trusight Cardio panel, comprising 174 genes associated with inherited cardiac conditions, were obtained from Imperial College London (London, UK; N=2150), the Aswan Heart Centre (Aswan, Egypt; N=1273), the Careggi University Hospital (Florence, Italy; N=219) and the Motol University Hospital (Prague, Czech Republic; N=132). These comprised 1041 patients affected with hypertrophic cardiomyopathy, 928 affected with dilated cardiomyopathy and 1805 unaffected volunteers (UVOL). The number of samples each center contributed per cohort is available in Supplementary Table 1.

### 2.2 Analyzed genes

The analysis was focused on genes targeted by the Illumina Trusight Cardio panel and classified as moderate-, strong- or definitive-evidence genes for DCM (N=18) and HCM (N=14, 9 of which shared with DCM) by the gene-disease validity curations performed by the ClinGen consortium^23,24^. Disease-specific gene sets were analyzed on DCM and HCM samples, while the full set of 23 genes was analyzed in UVOLs. *ACTC1, ACTN2, JPH2, MYH7, PLN, TPM1, TNNC1, TNNI3* and *TNNT2* were analyzed in both DCM and HCM cohorts, while *BAG3, DES, DSP, LMNA, NEXN, RBM20, SCN5A, TTN* and *VCL* were analyzed only in DCM and *CSRP3, MYL2, MYL3, MYBPC3* and *TRIM63* were analyzed only in HCM.

### 2.3 Variant identification and data processing

All samples were analyzed with four methods to detect large (≥20bp) variants, relying on orthogonal computational approaches and representing optimal choices for identifying ≥20bp variants from short-read paired-end targeted sequencing on a set of highly correlated samples − given the use of the same sequencing method and panel − like the one analyzed here:

- the pangenome-based Velsera GRAF pipeline – for this pipeline two disease-specific pangenomes (for DCM and HCM) were created upstream, with the aim of maximizing the accuracy of large variant detection. Details concerning the pangenome creation are listed in paragraph 2.3.
- GATK HaplotypeCaller v4.1 (GATKhc); based on local reassembly - while not being a large variant caller per se, GATKhc is currently among the optimal choices for detecting variants in the 20bp-50bp range at read depths >50x^25^, and is widely used in both the research and the diagnostic settings. Data processing and variant calling were performed following the Broad Institute’s Best Practices for germline short variant identification, utilizing BWA v0.7.17 for aligning reads to the GRCh38 reference genome. The same version of BWA was used also prior to calling variants with Manta and ExomeDepth (see below).
- Manta v1.6.0; large-variant caller combining paired-end and split-read evidence, developed by Illumina and optimized for detecting large variants from short-read paired-end data^26^.
- ExomeDepth v1.1.16 (ED); read-depth based copy number variant caller, optimized for analyzing highly correlated samples^27^. Since ED compares the analyzed samples to a set of reference samples, UVOLs were used as reference set when analyzing DCM and HCM patients. In the analysis of UVOLs, the cohort was split in half, performing the variant calling on 902 individuals and utilizing the remaining 903 as reference samples. The 902 UVOLs that underwent variant calling comprised those in which ≥1 variant of interest was detected by any other caller, for comparative purposes. Given the substantially higher number of variants detected by ED in comparison with the other callers, the Bayes Factor calculated by ED and reflective of the confidence of each variant call was utilized to filter variants to a manageable shortlist of candidates for lab-based validation, excluding from further analysis those variants with a Bayes Factor below 26.3 (see Paragraph 2.5 for details).

### 2.4 Variant prioritization

Variants ≥20bp in size in the genes of interest identified by the 4 tools were filtered further according to the following criteria:

- PASS filter (i.e. all caller-specific quality control steps passed) for variants called by GRAF, GATKhc and Manta, or with BF≥26.3 for those called by ED (see paragraphs 2.3 and 2.6 for details).
- Predicted protein-altering effect.
- If in *TTN*, variants altering exons constitutively expressed in the heart (spliced into >90% of the cardiac transcripts, Percent Spliced In >90%)^28^.

In the downstream analysis prior to variant validation, distinct variant calls were considered relative to the same event if:

- They were made in the same sample and in the same gene
- The overlap between the two variants was ≥50% for both variants

### 2.5 Creation of the disease-specific pangenomes

The procedure for the creation of the pangenomes is displayed in Figure 1. For each of the two pangenomes (HCM- and DCM-specific), the following steps were carried out:

- An initial pangenome reference was created by augmenting GRCh38 reference sequence with alternative contigs and high-confidence variants selected from the results reported by the 1000 Genomes Phase 3 study^29^, the Simons Genome Diversity Project^30^ and the indels database by Mills et al.^31^.
- The Velsera GRAF Aligner (GRAL) v1.2 was used to align raw sequencing data from all 3774 samples to this initial pangenome.
- Manta v1.6.0 was used to perform an initial round of large variant calling on all samples aligned to the pangenome. This was done because augmenting the pangenome with a “raw” set of large variants called by another tool significantly increases GRAF’s abitity to identify these variants downstream. Additional details concerning this step are provided in the Supplementary Tables 2 and 3. High quality, sequence resolved variants called by Manta were added to the pangenome reference. This was done by means of the Velsera Graph Construction Pipeline, displayed in Figure 1 and previously described^14^. Here, two distinct sets of variants were added to the pangenome, to create two disease-specific pangenomes:

`o` The DCM-specific pangenome was created by adding variants detected in both DCM and/or UVOL samples.
`o` The HCM-specific pangenome was created the same way, adding variants found in HCM and/or UVOL samples.

**Figure 1.**
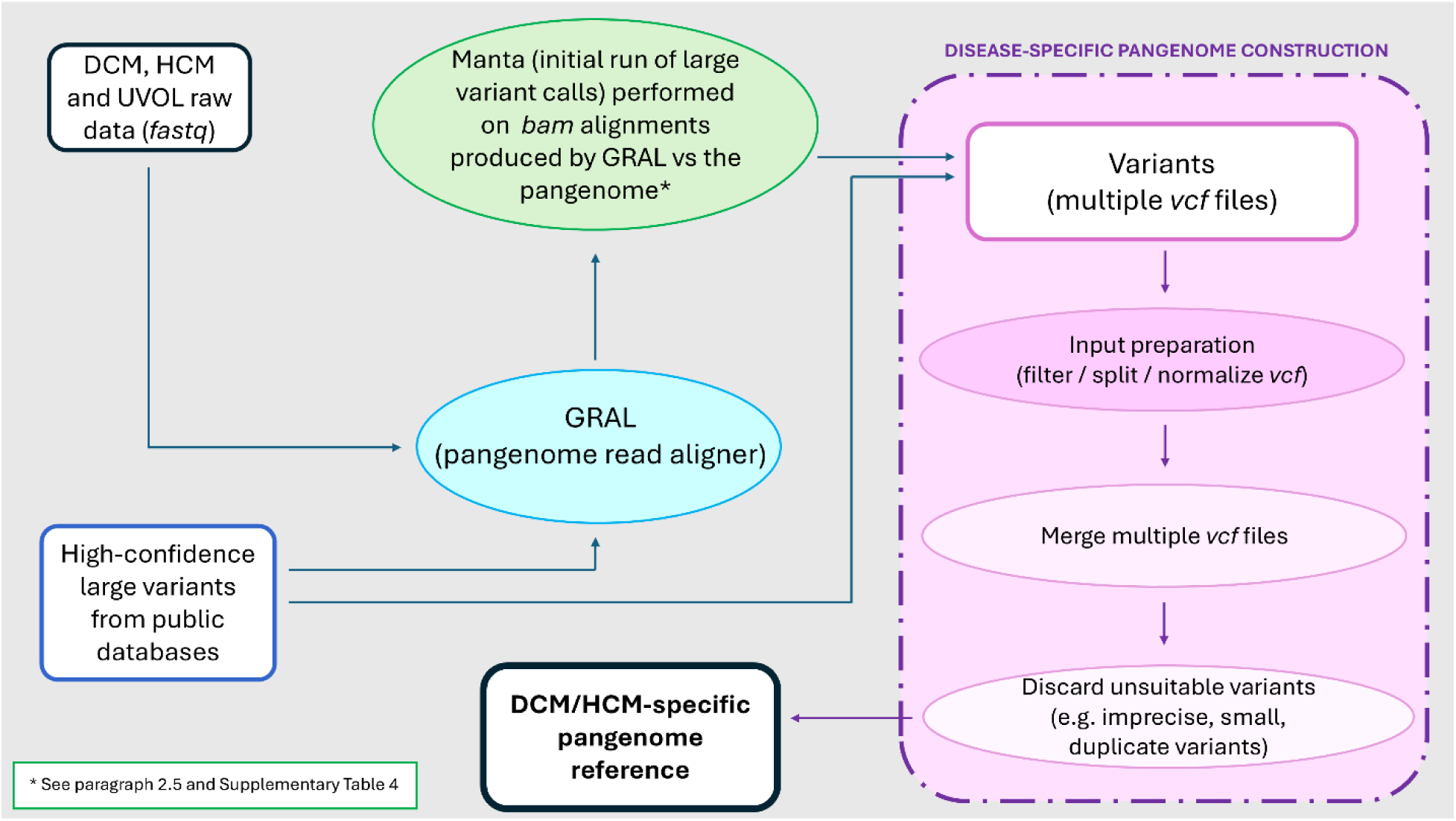
disease-specific pangenome building workflow.

In performing the core analysis on the 3,774 samples, the Velsera GRAF Germline Variant Calling Pipeline v1.2 was applied separately on the DCM and HCM samples using the respective DCM and HCM large-variants-enriched pangenomes, while UVOLs were analyzed with both. Commands executed in running the GRAF pipeline are listed in Supplementary Table 4.

### 2.6. Derivation of an empirical custom Bayes Factor cutoff for ExomeDepth variant calls

ED generally favors sensitivity over specificity and consequently tends to call a high number of variants. Each variant is scored with a Bayes Factor (BF) reflective of ED’s confidence in the variant call, corresponding to the log10 likelihood ratio of the presence of the variant over the null hypothesis (its absence). Aiming at a manageable number of candidate variants to bring forward for lab-based validation while avoiding to exclude potential true positives, we derived an empirical BF cutoff based on the burden of large variants (in the 32 analyzed genes) in the structural variants dataset of the Genome Aggregation Database (gnomAD SV v2.1, comprising 10,738 WGS samples and reporting variants >50bp in size). Initial calls made by ED comprised 71 distinct variants >50bp in 102 UVOLs (102 carriers in 902 analyzed UVOLs; 11.3%). We computed the burden of >50bp variants in the 32 analyzed genes in gnomAD SV (24 of 10,738 carriers; 0.22%) and, aiming at being conservative, we applied a BF threshold yielding a burden of 1% (∼ 4.5-fold the gnomAD burden) of these variants in UVOLs (9 carriers in 902 UVOLs; BF≥26.3).

### 2.7 Lab-based variant validation

Variants detected by one or more of the 4 tools and that passed the described filtering steps (N=36) were brought forward for lab-based validation. Validation was performed by means of one or both of the following:

- PCR, used as a first-line assay to investigate the presence of variants.
- Sanger sequencing, which was directly used to sequence the variant through its entire length when smaller than ∼500bp in size or to sequence the breakpoint regions of longer variants, whenever PCR provided evidence in favor of their presence.

For duplications, PCR primers were designed also considering the possibility of a duplication coupled with an inversion event. In the analysis of validation results, a variant call was considered correct (i.e. a true positive) if the breakpoints – and the inserted sequence, in case of insertions – were precisely identified with no imprecisions by the variant caller.

Genomic regions of interest were PCR amplified by using optimized genomic primers (available upon request). These primers were confirmed to lack known variants using the gnomAD database. PCR products were purified with ExoSAP-IT followed by bidirectional sequencing with the BigDye Terminator v1.1 Cycle Sequencing kit on the SeqStudio™ Genetic Analyzer System. Sequences analysis was performed with the Sequencher 5.0 software, and variants were annotated according to the Human Genome Variation Society nomenclature using the Alamut Visual Plus version v1.11 (SOPHiA GENETICS).

### 2.8 Estimation of the false negative variant calling rate

Running the 4 tools on the analyzed cohorts and validating all computationally predicted variants in the lab is informative about the tools’ precision, but not about their recall (given the impossibility to quantify real variants gone undetected by all tools, i.e. false negative variant calls). For this reason, we ran GRAF, GATKhc, Manta and ExomeDepth on whole exome sequencing (WES) short reads generated from the standard HG002 Genome In A Bottle (GIAB) sample using the Agilent v5 exome panel, and compared the set of called variants with the benchmark set published by the Human Genome Structural Variation Consortium^32,33^.

## 3. RESULTS

### 3.1 Prioritized variants

Following the filtering steps outlined in paragraph 2.4, a total of 39 sample-variant calls made by the 4 tools were prioritized for further analysis (Table 1 and Supplementary Table 5). ExomeDepth is the tool that made the highest number of variant calls (N=24, 62% of the total), followed by Manta (9, 23%), GATKhc (3, 8%) and GRAF (3, 8%). Of note, the application of the custom filter applied to ExomeDepth calls described in paragraph 2.6, which excluded variants with BF<26.3, caused the exclusion of 217 additional variant calls made by the tool across the three cohorts. As far as GRAF variant calls are concerned, no difference in terms of prioritized variants was observed in the UVOL cohort when aligning against the HCM or the DCM-specific pangenome.

**Table 1.**
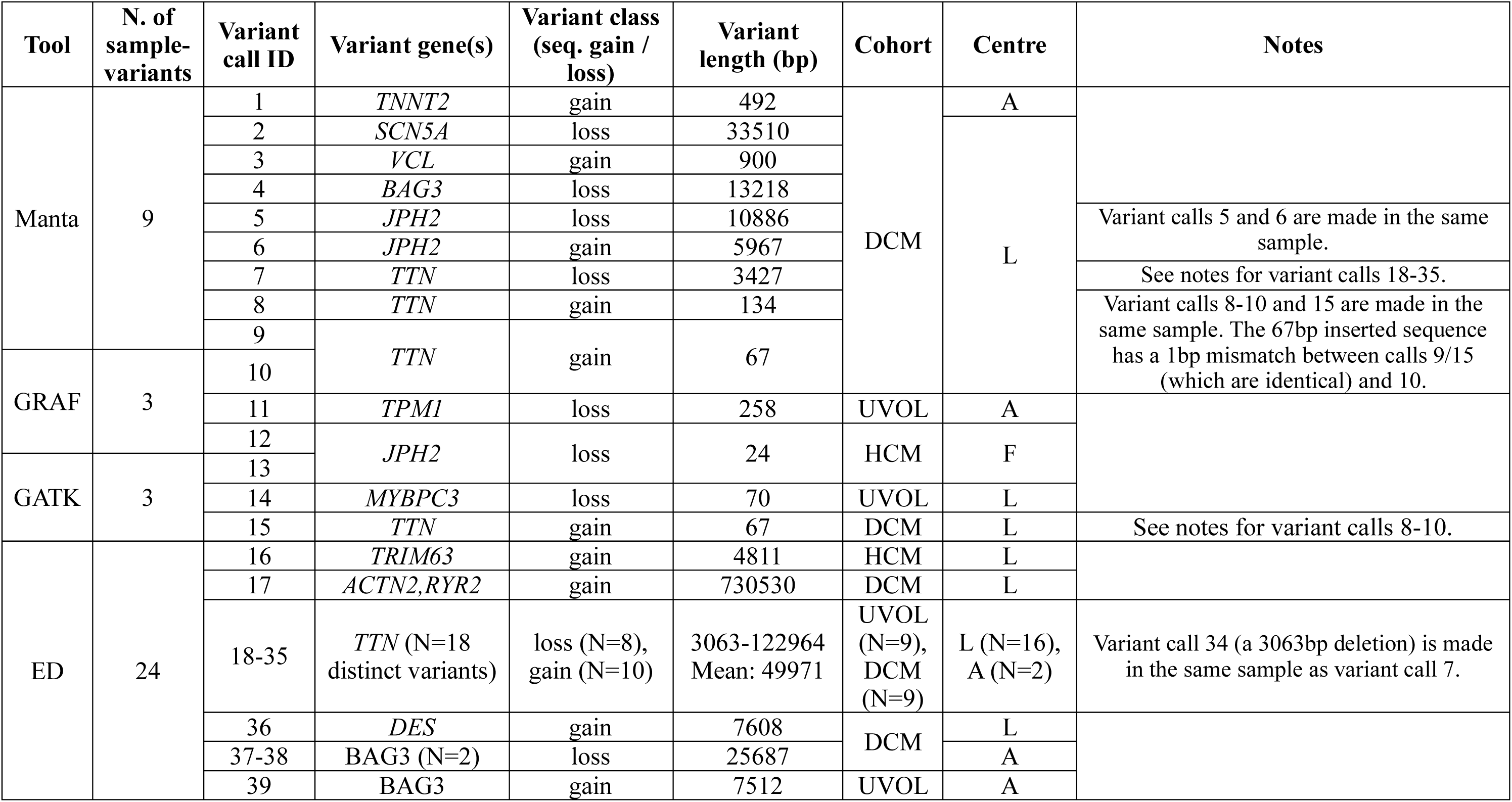
Variants prioritized for lab-based validation following the filtering steps detailed in Paragraph 2.4. Further details of each variant (including exact predicted breakpoints, sequence (for variants shorter than 1000bp), annotations and exons involved) are provided in Supplementary Table 5. L=London, A=Aswan, F=Florence.

These 39 initially prioritized variant calls were subsequently mapped to 34 distinct mutational events (Figure 2), as per the criteria outlined in Methods (paragraph 2.4). Variants called by different tools were characterized by distinct variant length ranges, with ED uniquely calling kb-range variants and, in contrast, GATKhc and GRAF making calls well below 1kb in size (Figure 2).

**Figure 2.**
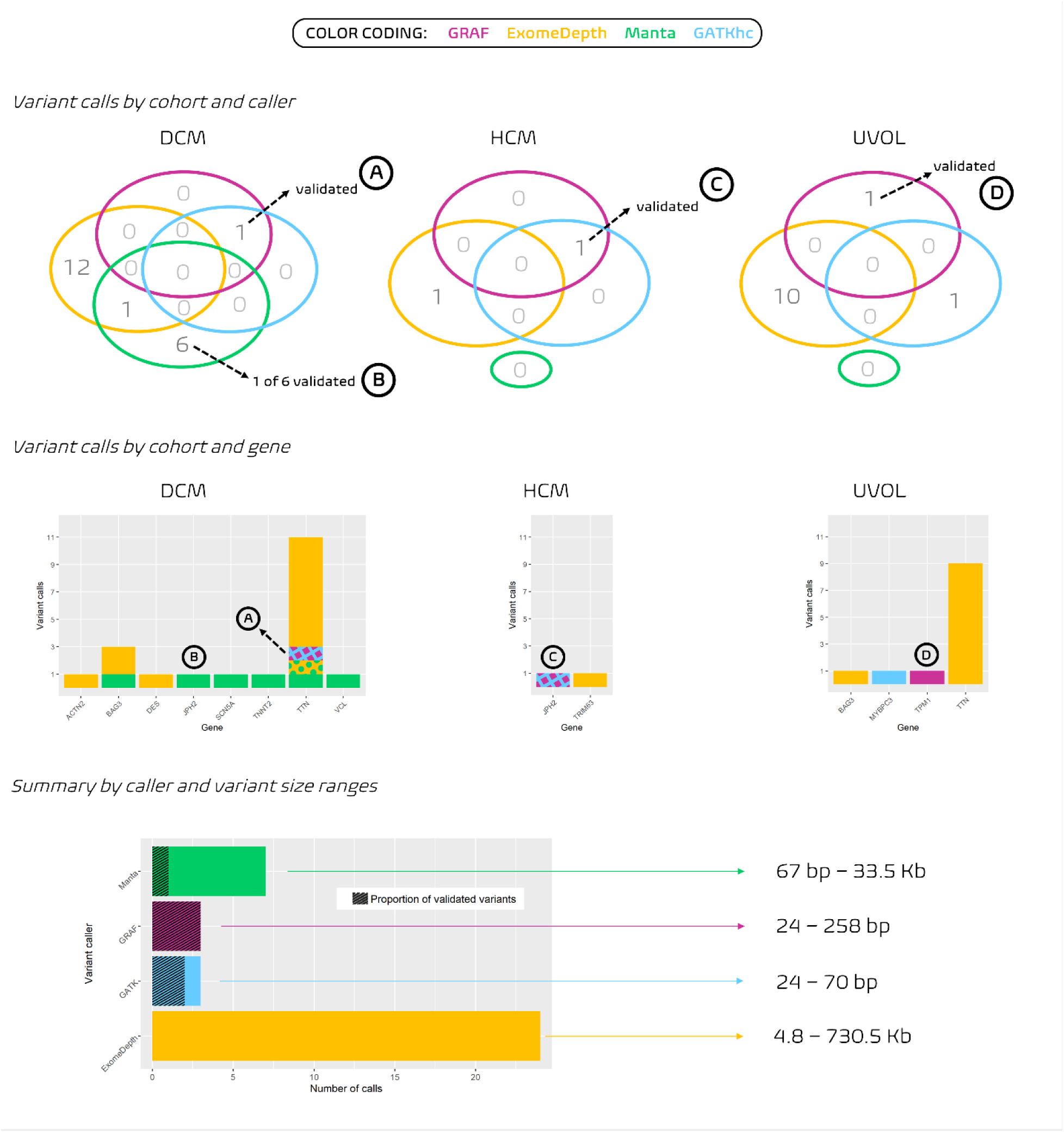
overview of the prioritized variant calls made by the 4 callers. The color coding outlined in the legend is consistent throughout the figure. Top: distribution of variant calls by cohort and by variant caller, alongside details on which variants were confirmed by Sanger and/or PCR in the lab (“validated”), each labeled with a letter A-D. Middle: Distribution of variant calls by cohort and by gene, with details concerning variant caller(s) (bi-color patterns indicate the two callers that made the call) and which variants were validated in the lab. Bottom: total number of prioritized calls made by each caller, alongside the proportion of variants confirmed by lab-based validation, and variant calls’ size range per caller.

### 3.2 Validated variants and burden of potentially pathogenic ≥20bp variants in DCM, HCM and in controls

Lab-based variant validation by means of Sanger sequencing and/or PCR (see Methods) confirmed the presence of 4 of the 34 prioritized variants (11.8%). In all 4 cases, the validated variants had been identified with precise breakpoints (and correct sequence, in case of insertions) by at least one tool. The 4 callers were characterized by markedly different precision (i.e. true positive calls over total calls), with GRAF achieving 100%, followed by GATKhc (66.7%), Manta (14.3%) and ED (0%). Details of the validated variants are provided in Table 2. Based on these results, we detected a burden of potentially pathogenic variants ≥20bp in size of 0.22% in DCM (1 in 928 samples), 0.1% in HCM (1 in 1041) and a background rate of 0.05% (1 in 1805) in unaffected controls.

**Table 2.** Details concerning the 4 variants validated in the lab. Column “Centre”: L=London, A=Aswan, F=Florence. Column “Validation strategy”: P=PCR, S=Sanger.

| Label in Fig.2 | Var. call ID(s) | Gene | Variant class | Variant length (bp) | Cohort | Centre | Variant caller(s) | Validation strategy | Predicted effect |
| --- | --- | --- | --- | --- | --- | --- | --- | --- | --- |
| Ⓐ | 10,15 | *TTN* | gain | 67 | DCM | L | GRAF, GATKhc | S | Frameshift variant |
| Ⓑ | 5-6 | *JPH2* | gain+loss (in *cis*) | 4919 (10886 del + 5967 dup) |  |  | Manta | P + S | Dup. of exon 3 and part of exon 6, fused together |
| Ⓒ | 12,13 | *JPH2* | loss | 24 | HCM | F | GRAF, GATKhc | S | Inframe deletion |
| Ⓓ | 11 | *TPM1* | loss | 258 | UVOL | A | GRAF | P + S | Intronic deletion comprising the splice donor site |

### 3.3 Estimation of the callers’ false negative rate

Of the 52 truth set variants ≥20bp in length spanning WES-targeted regions of sample HG002, GRAF was the tool that detected the most (n=35, false negative rate 32.7%), followed by GATKhc (n=26, 50.0%), Manta (n=1, 98.1%) and ED (n=0, 100.0%).

## DISCUSSION

In this study, we processed a total of 3774 technically matched, short-read panel-sequenced cardiomyopathy patients and controls with a pangenome-based (GRAF) and three reference-based orthogonal approaches (GATKhc, Manta and ED) to detect variants ≥20bp in size. Of note, each of these approaches was selected because theoretically optimal for processing these data, as detailed in paragraph 2.3.

The underlying rationale was, firstly, that of evaluating whether the adoption of pangenomes can aid detection of large (especially if longer than the sequencing read length) insertions and deletions from targeted sequencing data, as gene panels and whole-exome sequencing are still the most widely used sequencing assays in the diagnostic context. Secondly, we were also interested in providing a burden estimate for potentially pathogenic variants ≥20bp in DCM- and HCM-, taking into account the background rate in the population of such variants in disease-associated genes.

Overall, the low validation rate characterizing the variants detected by the four callers (4 of 34 variants, 11.8%) highlighted once more the intrinsic difficulty in bioinformatically predicting the presence of large variants from short sequencing reads. In our case, however, the four tools applied here were characterized by strikingly different variant validation rates, ranging from 0% (ED, 0 of 24 variants confirmed) to 100% (GRAF, 3 of 3 variants confirmed).

Of note, the initial number of calls made by ED had already been substantially reduced up front, considering that the application of the empirically derived BF-based cutoff described in paragraph 2.6 caused the exclusion of 90% variants prior to validation (217 of 241). While a (relatively) high number of false positive calls by ED was somewhat expected, considering that ED generally favors sensitivity over specificity, ED’s failure in detecting all the 4 validated variants in our study (absent also among the 217 discarded up front) compared with significantly better results obtained in previously published analyses can be probably explained, at least in part, by a combination of multiple factors. These range from the adoption of post-processing tools to further filter variants of potential diagnostic relevance prior to validation (e.g. ClassifyCNV^34^), to the specific processing of samples from the same sequencing run (to maximize the read depth correlation – although ED should automatically select the most highly correlated control samples). In addition, the vast majority of lab-validated variants detected by ED in these studies are very large (spanning from several kilobases to megabases in size) and that these variants are most likely completely absent in our cohorts^35,36^. In addition, the usage of panel data – as opposed to WES – may provide ED with a narrower, hence less informative, set of genes on which to compute read-depth correlation between cases and controls, although studies applying ED on gene panel data were previously published^37,38^.

GATKhc had a validation rate of 66.7%, with 2 of 3 identified variants validated by PCR and Sanger sequencing. Despite the very limited number of variants to draw conclusions, the size of these two variants (24bp and 67bp) compared with that of validated variants identified uniquely by other callers (258bp and ∼5kb) confirms GATKhc’s accuracy in identifying variants shorter than read length and especially with sizes in the lower end of the ≥20bp range^25^. Two things must be noted here – first, the 67bp insertion in *TTN* called by GATKhc (variant call #15 in Table 1 and Table 2) went undetected by GATKhc as we applied it in this work, but was identified by the in-house GATKhc pipeline in use at the London group, with a correct insert sequence and size (variant call identical to #10 made by GRAF). Aiming at not penalizing GATKhc, we considered this variant as called also by GATKhc throughout this work. Second, Manta also made similar calls at the same genomic location and in the same sample (i.e. variant call #8, with correct insert sequence and location but wrong ploidy/insert size and variant call #9, with correct ploidy/insert size and location, but incorrect sequence). While the mismatch between the real insert sequence (correctly recognized by GRAF and GATKhc) and the one detected by Manta was minimal (a 1bp, G/T mismatch at the 26^th^ inserted base, as shown in Supplementary Table 5), we did not consider variant call #9 correct given the key importance of identifying genetic variants correctly and unambiguously in the diagnostic setting. Details concerning the lab-based validation of these two variants are provided in Supplementary Figures 1 and 2.

On the other hand, Manta (characterized by an overall validation rate of 14.3%, as shown in Figure 2) was the only tool that identified a complex, kb-range duplication/deletion *JPH2* event (variant calls #5 and #6 in Table 1 and Table 2), remarkably with correct breakpoints. This large variant is the result of a 5967bp duplication spanning part of exon 6 (in the 3’ untranslated region (UTR)), exon 5, exon 4 and part of exon 3 of *JPH2* combined with a larger 10886bp deletion, spanning a larger portion of the 3’ UTR and again exons 5, 4 and part of exon 3. Lab-based validation confirmed these two variants in *cis* as part of a unique complex event, with the net result being a loss of 4919bp but also involving the creation of a chimeric 1997bp sequence formed by parts of UTR exon 6, intron 2 and exon 3 (details in Supplementary Figure 3).

Notably, this complex *JPH2* variant is the only validated variant missed by GRAF. While this shows how pangenome-based approaches like GRAF should be improved further before being used as an all-in-one solution in the diagnostic setting, the presence of the 11kb deletion in particular was supported by a remarkably minor proportion of reads (3 of 23 paired reads and 12 of 141 split reads). The current GRAF implementation does not resolve this complex event because it works by looking for the best unique mapping of each read, even when this requires clipping the read ends. This strategy is generally optimal for most variants (regardless of their size) but may be penalized when such a marginal share of the total reads supports the presence of the variant. In contrast, tools primarily relying on split-read evidence like Manta may be favored yet at the cost of making more false positive calls. However, the sensitivity of split-read methods like Manta in cases like this (i.e. variants supported by a very low number of reads at the boundaries of the target region) is one of the reasons why we advise populating the initial pangenome with variants called by a tool of this kind, as we did here (see paragraph 2.5). Regardless, GRAF is the only tool we applied here that did not make any false positive large variant call across the 3774 samples we processed and is it notably the only variant caller that detected a validated *TPM1* 258bp deletion in an unaffected individual (variant call #11). This is an intronic deletion including the last base of *TPM1* exon 5 and its splice donor site and, while also this variant is at the boundary of the target region, GRAF realignment revealed a strong support in favor of its presence (336 reads of 607). The important improvement in terms of read alignment accuracy achieved through the usage of the pangenome (as opposed to the conventional linear genome) is demonstrated also by the fact that this *TPM1* variant was detected also by Manta when run on the pangenome alignment (Supplementary Table 3) and was among those added to the pangenome itself prior to running GRAF. Conversely, Manta did not detect this variant when conventionally run on the .*bam* files produced by BWA, with reads aligned against GRCh38. Details concerning the wet lab-validation of this variant are provided in Supplementary Figure 4.

Overall, GRAF is the approach that achieved the highest precision (i.e. the validation rate, see paragraph 3.2) and, assuming the absence of additional variants in the genes of interest gone undetected by all tools, also the highest recall (75%, followed by GATKhc (50%), Manta (25%) and ED (0%)). The F1 score, summarizing each tool’s performance, was 0.86 for GRAF, 0.57 for GATKhc, 0.18 for Manta and 0 for ED.

An approximate estimate of how many variants may have been missed by all four tools can be made based on the results obtained on GIAB sample HG002, since this sample was sequenced with WES using paired-end short-read sequencing, and its variants are known. On this WES sample, GRAF and GATKhc were characterized by false negative rates of 32.7% and 50%, which are close to those we observed on our cohorts treating the 4 validated variants as the truth set (25% and 50%, respectively). ED was characterized by a WES-wide false negative rate of 100% on this sample but – while we provided ED with a pool of 22 WES control samples from paired-end, short-read sequencing to choose from – these were not sequenced in the same center and the read depth correlation between HG002 and the selected control was 0.73, below the minimum of 0.97 suggested for ED to produce reliable results. However, the mean (±SD) correlation observed when processing the HCM, DCM and UVOL panel samples was 0.9958±0.0177. Manta’s false negative rate in HG002 (98%) is further from the one we observed on our cohorts (75%), though this is entirely due to the complex *JPH2* variant uniquely identified by Manta. While these observations are based on the extremely limited number of validated variants in the analyzed cohorts, they are not suggestive of a likely presence of additional large variants that went undetected in this analysis. Based on this, we estimate the burden of potentially pathogenic large variants in DCM and HCM to be very low (≤0.2% for both HCM and DCM in this analysis). Limiting the comparison to the same genes screened here, a recently published large variant study in DCM and HCM (variant size-matched and featuring the largest cardiomyopathy cohorts surveyed for CNVs to date) detected comparable burdens to those observed in our cohorts (11/3550 (0.3%) vs 2/928 (0.2%) in DCM, Fisher test p=1 and 13/6779 (0.2%) vs 1/1041 (0.1%) in HCM, Fisher test p=1)^39^, further supporting the lack of additional variants gone undetected in samples analyzed here. In addition, the validated *TPM1* deletion detected in an unaffected individual may be reflective of the background rate of these variants in the population and – even if negligible – may translate into an even lower proportion of cardiomyopathy cases explained by large variants. Of note, however, no other potentially pathogenic variants (including SNVs and small indels) were identified in the three carrier patients detected here, in whom these variants could be directly causative.

In conclusion, this work took cardiomyopathies as a use case to investigate the impact of pangenome-based approaches in detecting large variants from short-read targeted sequencing, constituting the most widely used approach in the diagnostic context. Results show that pangenome approaches like GRAF – while still not achieving total recall – outperform orthogonal reference-based methods. Findings also reflect how reliably identifying these variants remains challenging: golden standard and highly precise tools developed to detect exclusively or primarily small variants (like GATKhc and AI-based DeepVariant^40^) don’t identify large variants, while less precise methods such as Manta and ExomeDepth accompany the large variant calls with high false positive rates, making validation impractical. The GRAF pangenome workflow, however, represents an optimal and versatile option for identifying all variant types (including small ones^14^), with the augmented pangenome providing scaffolds for correct mapping of the sequencing reads. The GRAF automated workflow is suitable for use by institutions with access to sequencing data from large cohorts to create disease specific pangenomes. We present this work in the hope that wider adoption of pangenome-based methods will improve the rate of positive diagnoses in patients suffering from genetic disorders, without imposing additional costs on healthcare providers.

## Data Availability

All data produced in the present study are available upon reasonable request to the authors

## ACKNOWLEDGEMENTS AND FUNDING

JSW was supported by Sir Jules Thorn Charitable Trust [21JTA], The Medical Research Council (UK), The British Heart Foundation [RE/18/4/34215, SP/17/11/32885, BBC/F/21/220106] and the NIHR Imperial College Biomedical Research Centre. PP, JB and MM were supported by the Ministry of Health of the Czech Republic (grant 15-34904A) and Conceptual Development of Research Organization, Motol University Hospital, Prague (grant 00064203). VC, VB, MC and MR extend sincere thanks to the Fazzo Cusan family for its generous support. We are also grateful to Dr. Beatrice Boschi and Dr. Irene Giotti from the Careggi University Hospital (Florence, Italy) for having provided the Sanger chromatogram displayed in Supplementary Figure 2. For the purpose of open access, the authors have applied a Creative Commons Attribution (CC BY) licence to any Author Accepted Manuscript version arising.

## ETHICAL APPROVAL

All patients gave written informed consent, and all studies were approved by the relevant regional research ethics committees, and adhered to the principles set out in the Declaration of Helsinki. For London samples, ethical approval was granted by the Hampshire B Research Ethics Committee (09/H0504/104+5 and 19/SC/0257) and by the London - West London & GTAC Research Ethics Committee (09/H0707/69). For Prague samples, approval was granted by the IRB of Motol University Hospital (Reference No. EK-323/23).

## SUPPLEMENTARY NOTES

### Data and code availability

All software required to reproduce the results we obtained with GATKhc, GRAF and Manta is made available as Docker images. The workflows are implemented as CWL scripts, and can be conveniently executed using cwl-runner (https://github.com/common-workflow-language/cwltool).

A small dataset is made available for testing the local install of the pipelines (`inputs_small_disease_sv_public.zip`). Once the cwl-the files in this archive have been extracted, the following commands will execute the workflows, automatically downloading and installing the required docker images:

`cwl-runner --debug --tmpdir-prefix $PWD/temp_folder/ --outdir`

`outputs_small_disease_sv_test`

`inputs_small_disease_sv_public/disease_sv_graph_building_workflow.json`

`inputs_small_disease_sv_public/small_disease_sv_input.yaml`

This command will run the SV calling workflows on the test samples and create BAM and VCF output file in the folder `outputs_small_disease_sv_test`.

The Docker images downloaded by the workflow are all hosted on the public Velsera Docker registry:

- images.sbgenomics.com/ozem_kalay/gwgs-u:1.2
- images.sbgenomics.com/ozem_kalay/htslib-1-14:0
- images.sbgenomics.com/ozem_kalay/resolve_sv:0
- images.sbgenomics.com/ozem_kalay/bcftools-1-15-1:2
- images.sbgenomics.com/ozem_kalay/manta-1-6-0:0
- images.sbgenomics.com/ozem_kalay/pangenome:0.12
- images.sbgenomics.com/ozem_kalay/gwgs-u:1.0rc3
- images.sbgenomics.com/ozem_kalay/rtg-tools:3.8.4
- images.sbgenomics.com/ozem_kalay/pangenome:0.12.1

The tools and methods presented in this study (with the exception of ExomeDepth) are available on all Velsera cloud platforms. The graph-based sequencing data analysis tools used in this study (Velsera GRAF) are freely available to all researchers on all Velsera academic cloud platforms (Cancer Genomics Cloud, Cavatica, BioData Catalyst). To access academic platforms, please reach out via the respective website. Restrictions apply for commercial use, please contact Velsera for terms and other details.

Scripts applied to filter raw GATKhc, GRAF and Manta variant calls, as well as to run ExomeDepth, and merge/harmonize results are available in GitHub repository https://github.com/fmazzarotto/pangenomes_panel_study.

## SUPPLEMENTARY FIGURES

**Supplementary Figure 1:**
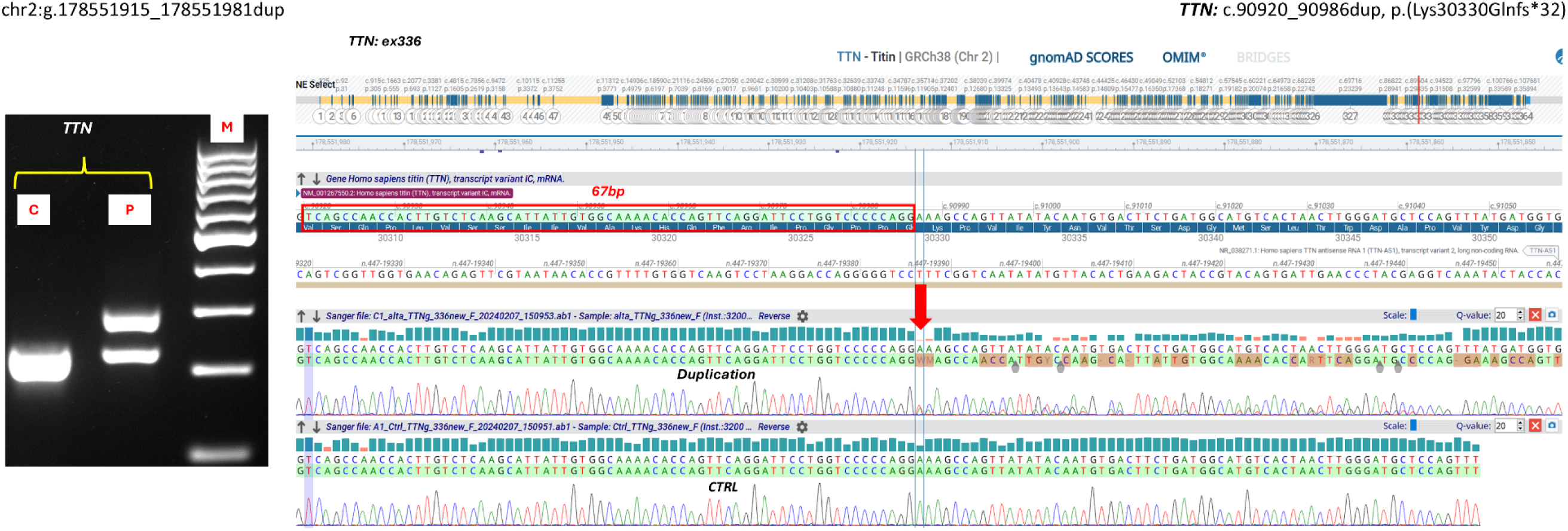
details concerning the PCR- and Sanger sequencing-based validation of variant Ⓐ (67bp duplication in TTN). In the PCR image on the left: C=control, P=patient.

**Supplementary Figure 2:**
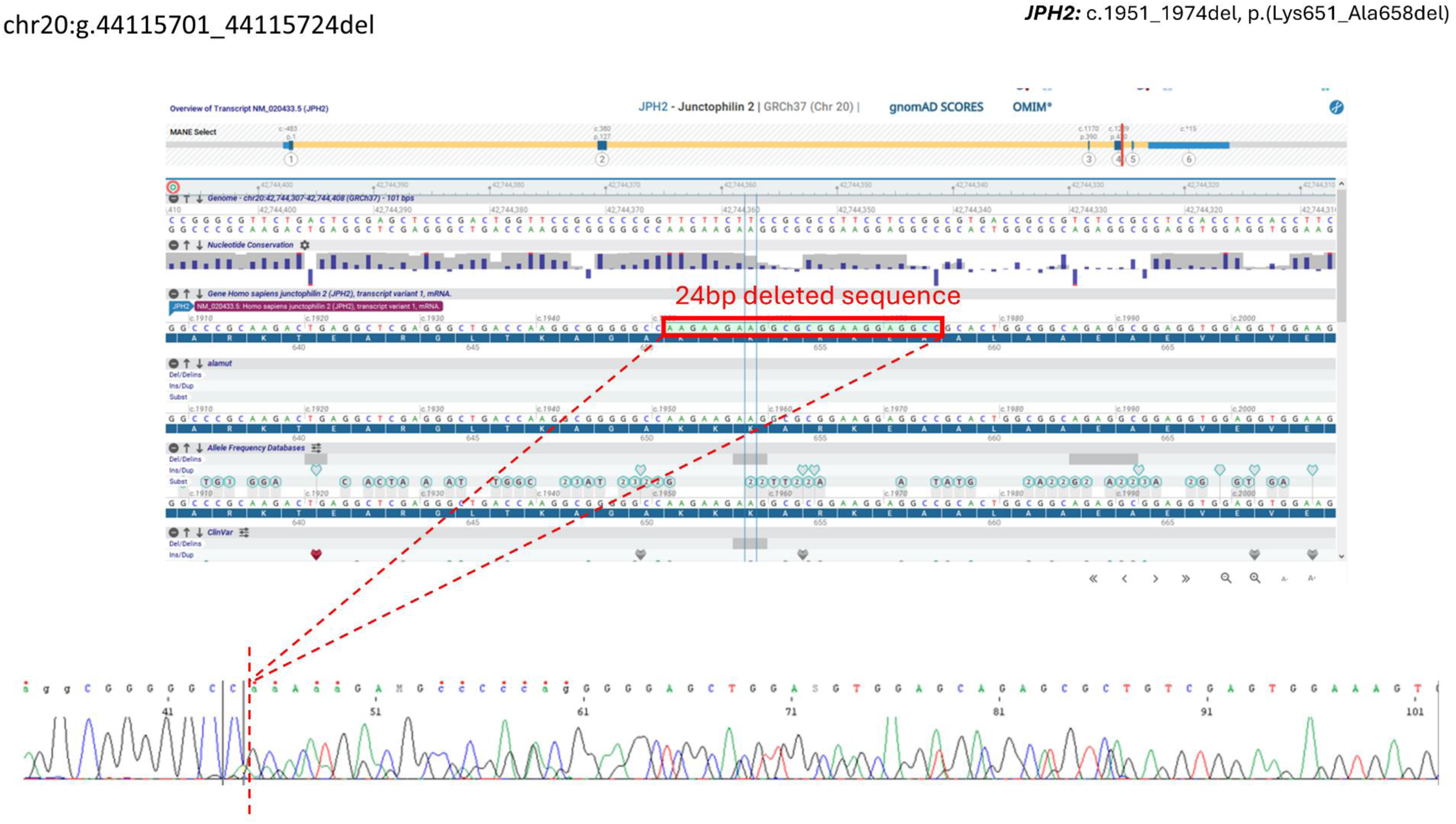
details concerning the Sanger validation of variant Ⓒ (24bp inframe deletion in JPH2).

**Supplementary Figure 3:**
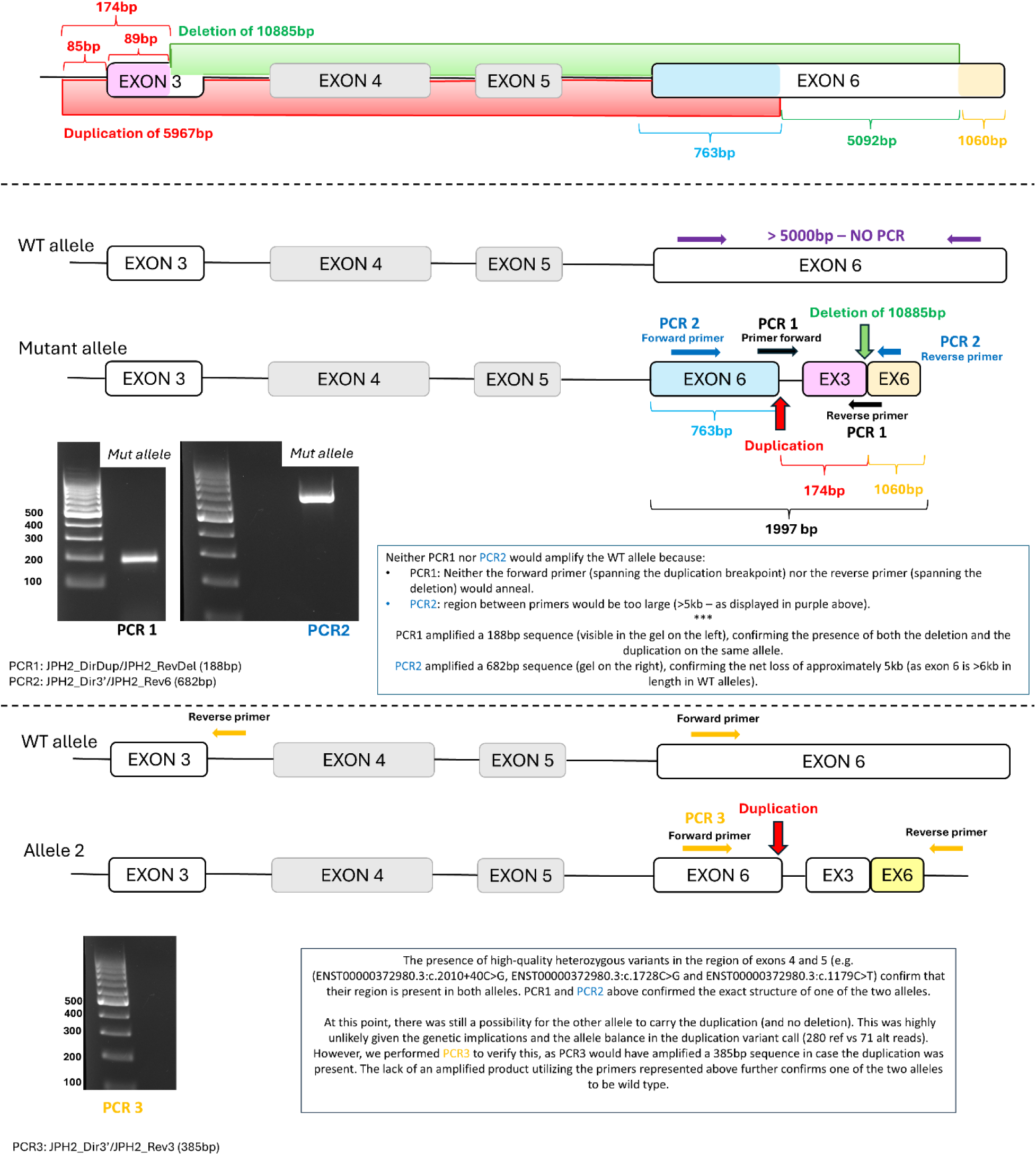
details concerning the three PCRs conducted in the lab to validate the presence of the large, complex JPH2 variant detected by Manta in a patient affected with dilated cardiomyopathy (variant Ⓑ). These results confirmed one allele to be wild-type, and the other allele to carry both events called by Manta (variant call IDs 5 and 6). The net result on the mutant allele is the loss of a 4919bp sequence, but also the creation of a chimeric sequence formed by a proximal part of UTR exon 6 (763bp), intron 2 (85bp), exon 3 (89bp) and a distal part of exon 6 (1060bp).

**Supplementary Figure 4:**
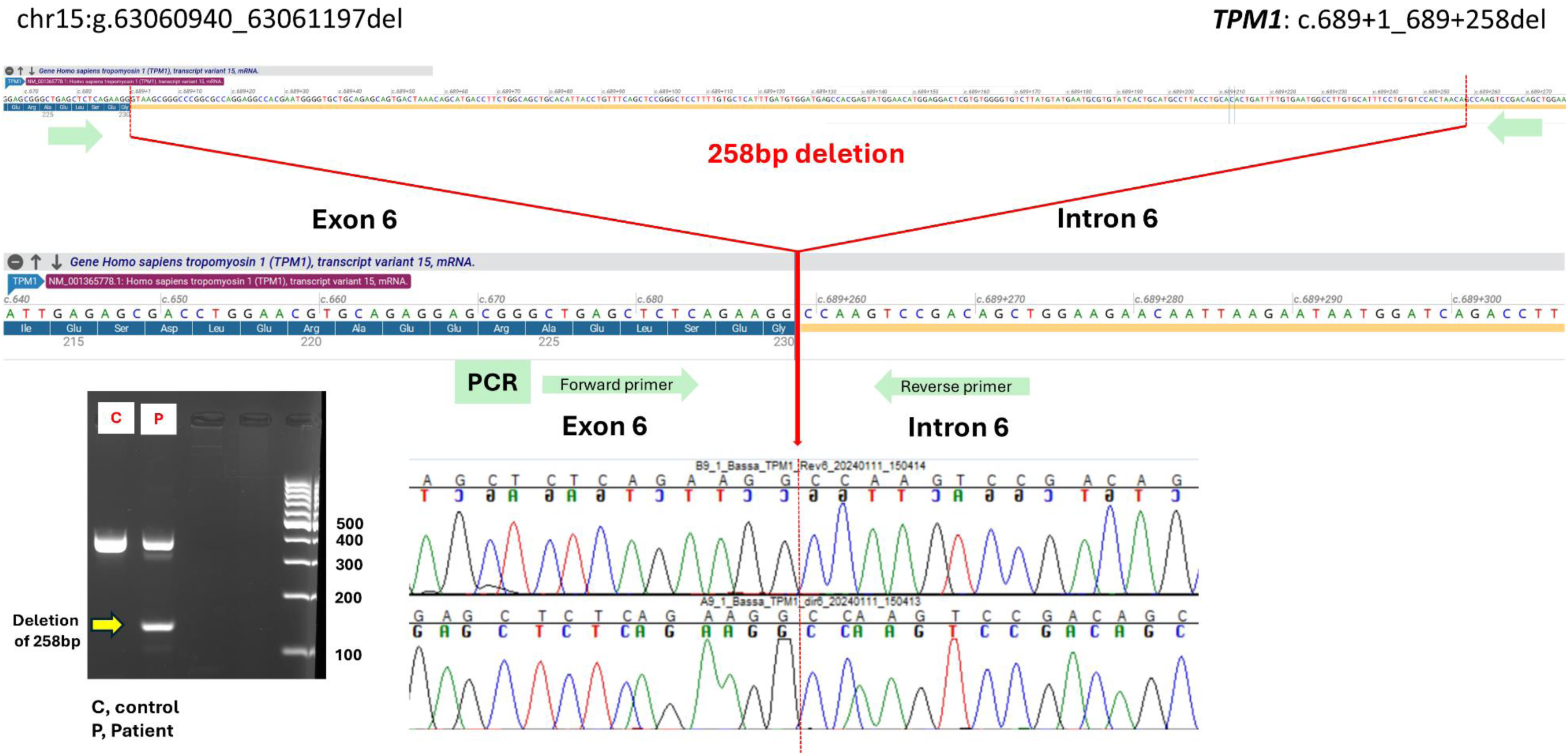
details concerning the lab-based validation of variant Ⓓ (258bp deletion in TPM1 detected in an unaffected volunteer).

## REFERENCES

1. Moreno-Cabrera, J. M. et al. Evaluation of CNV detection tools for NGS panel data in genetic diagnostics. Eur J Hum Genet 28, 1645–1655 (2020).

2. Teo, S. M., Pawitan, Y., Ku, C. S., Chia, K. S. & Salim, A. Statistical challenges associated with detecting copy number variations with next-generation sequencing. Bioinformatics 28, 2711–2718 (2012).

3. Schwarze, K. et al. The complete costs of genome sequencing: a microcosting study in cancer and rare diseases from a single center in the United Kingdom. Genetics in Medicine 2019 22:1 22, 85–94 (2019).

4. Wilke, M. V. M. B. et al. Diagnostic yield of exome and genome sequencing after non-diagnostic multi-gene panels in patients with single-system diseases. Orphanet J Rare Dis 19, 1–9 (2024).

5. Marwaha, S., Knowles, J. W. & Ashley, E. A. A guide for the diagnosis of rare and undiagnosed disease: beyond the exome. Genome Med 14, (2022).

6. Mazzarotto, F., Olivotto, I. & Walsh, R. Advantages and Perils of Clinical Whole-Exome and Whole-Genome Sequencing in Cardiomyopathy. Cardiovasc Drugs Ther 34, 241–253 (2020).

7. Gabrielaite, M. et al. A comparison of tools for copy-number variation detection in germline whole exome and whole genome sequencing data. Cancers (Basel) 13, (2021).

8. Gordeeva, V. et al. Benchmarking germline CNV calling tools from exome sequencing data. Scientific Reports 2021 11:1 11, 1–11 (2021).

9. Zhao, M., Wang, Q., Wang, Q., Jia, P. & Zhao, Z. Computational tools for copy number variation (CNV) detection using next-generation sequencing data: Features and perspectives. BMC Bioinformatics 14, 1–16 (2013).

10. Välipakka, S. et al. Improving Copy Number Variant Detection from Sequencing Data with a Combination of Programs and a Predictive Model. J Mol Diagn 22, 40–49 (2020).

11. Barcelona-Cabeza, R., Sanseverino, W. & Aiese Cigliano, R. isoCNV: in silico optimization of copy number variant detection from targeted or exome sequencing data. BMC Bioinformatics 22, (2021).

12. Danecek, P. et al. Detection and characterization of copy-number variants from exome sequencing in the DDD study. Genetics in Medicine Open 2, 101818 (2024).

13. Ameur, A. Goodbye reference, hello genome graphs. Nature Biotechnology 2019 37:8 37, 866–868 (2019).

14. Tetikol, H. S. et al. Pan-African genome demonstrates how population-specific genome graphs improve high-throughput sequencing data analysis. Nature Communications 2022 13:1 13, 1–11 (2022).

15. Ebler, J. et al. Pangenome-based genome inference allows efficient and accurate genotyping across a wide spectrum of variant classes. Nat Genet 54, 518 (2022).

16. Wang, T. et al. Pan-genome analysis of 13 Malus accessions reveals structural and sequence variations associated with fruit traits. Nature Communications 2023 14:1 14, 1–15 (2023).

17. Groza, C. et al. Pangenome graphs improve the analysis of structural variants in rare genetic diseases. Nature Communications 2024 15:1 15, 1–12 (2024).

18. Pua, C. J. et al. Development of a Comprehensive Sequencing Assay for Inherited Cardiac Condition Genes. J Cardiovasc Transl Res 9, 3–11 (2016).

19. Rakocevic, G. et al. Fast and accurate genomic analyses using genome graphs. Nat Genet 51, 354–362 (2019).

20. Ceyhan-Birsoy, O. et al. Next generation sequencing-based copy number analysis reveals low prevalence of deletions and duplications in 46 genes associated with genetic cardiomyopathies. Mol Genet Genomic Med 4, 143–151 (2015).

21. Singer, E. S. et al. Characterization of clinically relevant copy-number variants from exomes of patients with inherited heart disease and unexplained sudden cardiac death. Genet Med 23, 86–93 (2021).

22. de Uña-Iglesias, D., Ochoa, J. P., Monserrat, L. & Barriales-Villa, R. Clinical Relevance of the Systematic Analysis of Copy Number Variants in the Genetic Study of Cardiomyopathies. Genes (Basel) 15, 774 (2024).

23. Ingles, J. et al. Evaluating the Clinical Validity of Hypertrophic Cardiomyopathy Genes. Circ Genom Precis Med 12, 57–64 (2019).

24. Jordan, E. et al. Evidence-Based Assessment of Genes in Dilated Cardiomyopathy. Circulation 144, 7–19 (2021).

25. Wang, N. et al. Tool evaluation for the detection of variably sized indels from next generation whole genome and targeted sequencing data. PLoS Comput Biol 18, (2022).

26. Chen, X., et al. Manta: rapid detection of structural variants and indels for germline and cancer sequencing applications. Bioinformatics 32, 1220–1222 (2016).

27. Plagnol, V. et al. A robust model for read count data in exome sequencing experiments and implications for copy number variant calling. Bioinformatics 28, 2747 (2012).

28. Roberts, A. M. et al. Integrated allelic, transcriptional, and phenomic dissection of the cardiac effects of titin truncations in health and disease. Sci Transl Med 7, (2015).

29. Fairley, S., Lowy-Gallego, E., Perry, E. & Flicek, P. The International Genome Sample Resource (IGSR) collection of open human genomic variation resources. Nucleic Acids Res 48, D941–D947 (2020).

30. Mallick, S. et al. The Simons Genome Diversity Project: 300 genomes from 142 diverse populations. Nature 538, 201–206 (2016).

31. Mills, R. E. et al. An initial map of insertion and deletion (INDEL) variation in the human genome. Genome Res 16, 1182–1190 (2006).

32. Ebert, P. et al. Haplotype-resolved diverse human genomes and integrated analysis of structural variation. Science 372, (2021).

33. Chaisson, M. J. P. et al. Multi-platform discovery of haplotype-resolved structural variation in human genomes. Nat Commun 10, (2019).

34. Gurbich, T. A. & Ilinsky, V. V. ClassifyCNV: a tool for clinical annotation of copy-number variants. Scientific Reports 2020 10:1 10, 1–7 (2020).

35. Royer-Bertrand, B. et al. CNV detection from exome sequencing data in routine diagnostics of rare genetic disorders: Opportunities and limitations. Genes (Basel) 12, 1427 (2021).

36. Tilemis, F. N. et al. Germline CNV Detection through Whole-Exome Sequencing (WES) Data Analysis Enhances Resolution of Rare Genetic Diseases. Genes (Basel) 14, 1490 (2023).

37. Cacheiro, P. et al. Evaluating the Calling Performance of a Rare Disease NGS Panel for Single Nucleotide and Copy Number Variants. Mol Diagn Ther 21, 303–313 (2017).

38. May, V. et al. ClearCNV: CNV calling from NGS panel data in the presence of ambiguity and noise. Bioinformatics 38, 3871–3876 (2022).

39. de Uña-Iglesias, D., Ochoa, J. P., Monserrat, L. & Barriales-Villa, R. Clinical Relevance of the Systematic Analysis of Copy Number Variants in the Genetic Study of Cardiomyopathies. Genes (Basel) 15, 774 (2024).

40. Poplin, R. et al. A universal SNP and small-indel variant caller using deep neural networks. Nature Biotechnology 2018 36:10 36, 983–987 (2018).

